# Latent Structure in EHR Data: Reconstruction of Diabetes Markers with Sparse NMF

**DOI:** 10.1101/2025.03.31.25324972

**Authors:** Ahmed Elhussein, George Hripcsak

## Abstract

The dimensionality of electronic health record (EHR) data continues to grow as more clinical variables are recorded, often resulting in redundancy, sparsity, and analytical intractability. In this study, we apply non-negative matrix factorization (NMF) to a high-dimensional laboratory dataset of patients with type II diabetes to estimate the minimum latent dimensionality required to preserve clinically meaningful information. Using both within-patient imputation and across-patient generalization tasks, we evaluate the ability of the learned representations to reconstruct two key clinical lab values: blood glucose and HbA1c. Our findings show that clinically acceptable accuracy can be achieved with a dimensionality reduction of up to 80% and a dimensionality of 230 to 300, supporting the presence of a compact, low-dimensional latent structure underlying high-dimensional clinical data.

## 1 Introduction

Electronic Health Records (EHR) now routinely capture a wide array of patient-level variables. While these rich datasets have driven many advances [11, 2], their dimensionality is expanding rapidly. Consequently, healthcare data often suffers from *the curse of dimensionality*: the exponential growth in required sample size as feature cardinality increases [4, 3]. This complicates the identification of meaningful patterns and relationships and makes analysis increasingly untractable.

While raw EHR data are high-dimensional, many variables are rarely independent, providing an opportunity for dimensionality reduction. This redundancy is embedded at multiple levels. At the physiological level, diseases typically affect multiple biological systems simultaneously, resulting in correlated changes across various measurements. For example, creatinine and urea nitrogen levels both reflect related aspects of renal function. At the clinical workflow level, protocols often involve standardized panels of tests that are ordered together, creating predictable co-occurrence patterns. For example, diabetes often involves concurrent measurements of glucose, HbA1c, and lipid profiles, generating correlated data points by design. Finally, the recording process itself introduces structural redundancy. For instance, diagnosis and lab codes often capture overlapping clinical concepts (*e*.*g*., “type 2 diabetes” vs. “diabetes with complications”). Together, these dependencies suggest that EHR data lie on a lower-dimensional manifold, where the effective dimensionality—the minimal number of latent factors required to explain the observed data—is substantially smaller than the raw feature count.

This inherent redundancy in clinical data can be leveraged to mitigate issues related to sparsity and noise that are common in EHR datasets. By leveraging relationships between observed features, it is possible to construct latent variables that capture and aggregate information across distinct but related features [10]. This enables reconstruction of more complete patient profiles, even in the presence of sparse data [6]. This approach also reduces noise, as the latent representations effectively average across multiple noisy measurements of related processes. By distilling high-dimensional features into efficient, noise-reduced representations that capture underlying clinical patterns, we can enhance both the completeness and reliability of patient data [13, 1].

Various dimensionality reduction techniques have been developed to exploit the underlying structure and correlations in EHR data. These methods aim to project high-dimensional observations into a lower-dimensional space while retaining key information. Classical approaches include Principal Component Analysis (PCA) which identifies orthogonal axes of maximal variance, and Non-negative Matrix Factorization (NMF), which decomposes data into interpretable additive components [5]. More recently, deep learning–based methods such as autoencoders have been used to capture nonlinear relationships [8].

Despite these methods, however, the intrinsic dimensionality, the minimal number of latent factors required to represent clinically meaningful patterns, of EHR data remains poorly characterized. In this study, we aim to estimate the range of number of latent dimensions necessary to retain clinically relevant information in high-dimensional EHR datasets. We apply NMF to a large patient cohort with type II diabetes. Using these methods, we explore how well clinically significant variables such as blood glucose and HbA1c can be reconstructed after dimensionality reduction, and quantify the extent of information loss across a range of factorization ranks.

## 2 Background

Medical data can be represented as feature matrix **X** ∈ ℝ^*N × M*^, with the *N* rows representing patients and the *M* columns representing clinical observations such as laboratory test results. Each element **X**_*i,j*_ contains the observed value for patient *i* on clinical feature *j*.

### 2.1 Non-negative Matrix Factorization

Matrix factorization (MF) is a dimensionality reduction technique frequently employed in medical data analysis to identify latent structures. Given a data matrix **X**, MF decomposes this matrix into the product of two lower-rank matrices **W** and **H** of dimensions *N* × *R* and *R* × *M*, respectively, such that:

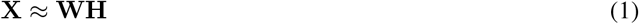

where *R* ≪ min(*N, M*) represents the reduced dimensionality. The factor matrices **W** and **H** capture a lower-dimensional representation of the original data, with each dimension potentially corresponding to a meaningful latent feature.

Non-negative matrix factorization (NMF) extends traditional MF by imposing non-negativity constraints on all elements of the factor matrices **W** and **H**. Formally:

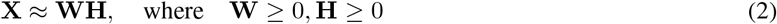

This non-negativity constraint enhances interpretability, as it ensures all factors can be understood as additive combinations of the underlying observed features. This aligns well with the nature of clinical data, where variables such as lab measurements or diagnosis codes are inherently non-negative. In this framework, each row of **W** can be interpreted as a patient’s expression across latent clinical concepts, while columns of **H** represent how original clinical features load onto those concepts.

**Sparse NMF** extends this framework by explicitly handling missing data. This is particularly valuable as most patients receive only a subset of all possible clinical measurements, often dictated by their conditions, care pathways, and encounter history etc. By limiting optimization to observed data, Sparse NMF produces more robust and generalizable representations, better reflecting real-world clinical heterogeneity.

While, standard NMF assumes that all entries of **X** are observed, Sparse NMF addresses this by introducing a binary masking matrix **M** ∈{0, 1} ^*N×M*^, where *M*_*ij*_ = 1 if *X*_*ij*_ is observed and *M*_*ij*_ = 0 otherwise. The optimization objective is then reformulated as:

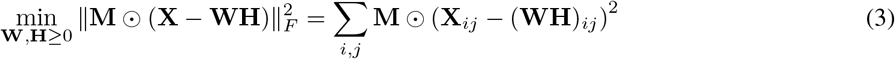

where denotes the Hadamard (element-wise) product. This formulation ensures that the reconstruction error is computed only over observed entries, avoiding distortion from unobserved or missing values.

## 3 Methods

### 3.1 Dataset and Preprocessing

#### Patient Cohort

We used the NewYork-Presbyterian Columbia University Irving Medical Center (NYP-CUIMC) electronic health record (EHR) dataset, which includes inpatient and outpatient clinical observations from 1995 to 2019. To define our study cohort, we identified adult patients (≥18 years) with type II diabetes, defined as having at least two encounters with diagnosis codes corresponding to descendants of ICD-9 code 250. Codes were included if their descriptions contained “type II” and excluded if they contained “type I”.

#### Lab Test Data

To construct a feature matrix, we focused on lab test results, given their ability to reflect both normal and abnormal physiological states. All available lab test observations for each patient were extracted. Given that the dataset spans multiple decades, lab test codes evolved over time. To address this, we harmonized lab test codes using a local terminology mapping tool `qrymed`.

For each patient, only the most recent observation for each lab test was retained to reduce temporal redundancy and limit sparsity. To further reduce computational burden, a random 50% sample of the eligible patient population was selected for analysis. All retained lab values were min-max normalized to prevent high-magnitude tests from dominating the latent space during model fitting. The final matrix consisted of 12,749 patients and 1,418 lab tests, with approximately 93% sparsity (see Table 1).

**Table 1:**
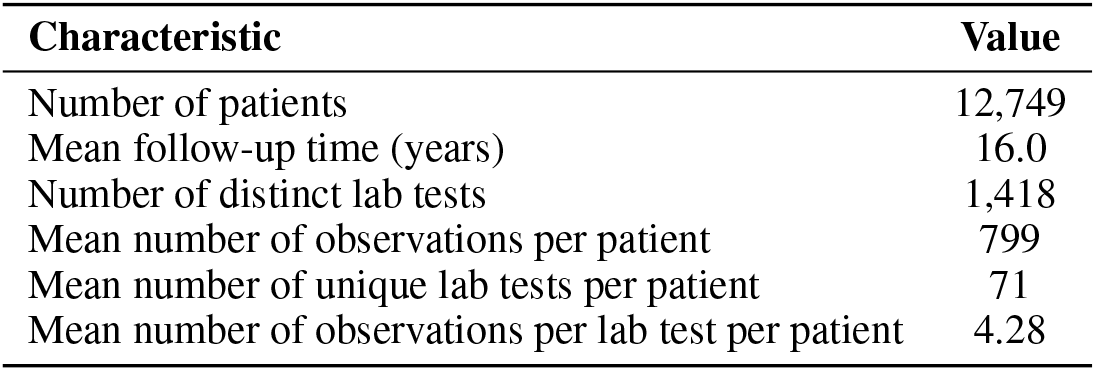
Summary statistics of the laboratory dataset.

### 3.2 Model Fitting

We applied a sparse-variant of NMF to the lab test feature matrix (equation 3). To identify the optimal latent dimensionality, models were fit across a broad range of latent dimensions: *k* ∈ {1, 5, 10, 20, 40, 60, 80, 100, 150, 200, 300, 400, 600, 800, 1000}. To better identify the optimal dimensionality, additional models were fit at finer rank intervals within ranges of interest, guided by trends in reconstruction error.

### 3.3 Evaluation approach

To assess the reconstruction fidelity of dimensionality-reduced datasets, we evaluated their capacity to recover two clinically actionable parameters in diabetes management: blood glucose (mg/dL) and glycated hemoglobin (HbA1c, %). We defined the optimal rank as the minimum dimensionality required to achieve clinically meaningful accuracy, defined by established thresholds: a mean absolute prediction error of *<* 10 mg/dL for blood glucose and *<* 0.5% for HbA1c. These thresholds are consistent with prior studies in clinical prediction modeling [7, 9, 12, 14].

We employed two distinct evaluation strategies:

1. **Masking Approach (within-patient prediction)**: We assessed the model’s ability to recover the value of a lab test when it was masked during training. Specifically, the target lab value (glucose or HbA1c) was removed from the matrix, and the model was trained using all other available lab results. The masked values were then reconstructed using the learned factor matrices. This approach assesses how well other lab variables capture information about the target variables, effectively treating the estimation as a missing data imputation task.
2. **Train-Test Split Approach (across-patient prediction)**: We tested whether the latent factors identified in one group of patients generalized to unseen individuals. The dataset was split into an 80% training set and a 20% test set. NMF was applied to the training set with all lab values available, and the learned feature matrix was used to reconstruct lab values for patients in the test set. This approach evaluates whether the latent factors identified in one patient group generalize effectively to unseen patients, testing the transferability and scalability of the identified factors.

These complementary evaluation strategies provide different insights: the first measures how much information about key clinical variables is embedded in other laboratory measurements, while the second assesses how well the learned data representations generalize to new patients.

## 4 Results

### 4.1 Dataset characteristics

#### Patient Cohort

A total of 21,261 patients with a diagnosis of type II diabetes were identified from the NYP-CUIMC EHR database. From this cohort, a random sample of 12,749 patients was selected for inclusion in the final analysis. The earliest recorded visit among these patients occurred in 1995 and the latest in 2019. The mean follow-up period—defined as the time between a patient’s first and last recorded visit—was 16.0 years.

#### Laboratory Data

The dataset contained 3,107 distinct laboratory tests. After harmonizing coding variations using `qrymed`, we consolidated these into 1,418 unique lab test features (45.6% of the original set). The final patient-by-lab matrix comprised 12,749 patients and 1,418 tests, with an overall sparsity of 93%.

Lab test utilization exhibited marked skewness: 100 tests accounted for 93.2% of all observations. Patients had, on average, 71 distinct tests with a 4.28 observations per test. 1,015 tests (71.6% of features) were observed in fewer than 1% of patients, reflecting substantial long-tail sparsity.

### 4.2 Evaluation

#### Masking approach

In the first evaluation we masked specific lab values during training and then predicted them from the latent representation. Figure 1 illustrates the mean prediction error across varying latent dimensions for blood glucose and HbA1c. Applying clinically meaningful thresholds we found that a minimum rank of ~ 230 was required for accurate glucose reconstruction, and a minimum rank of ~ 280 for HbA1c.

**Figure 1:**
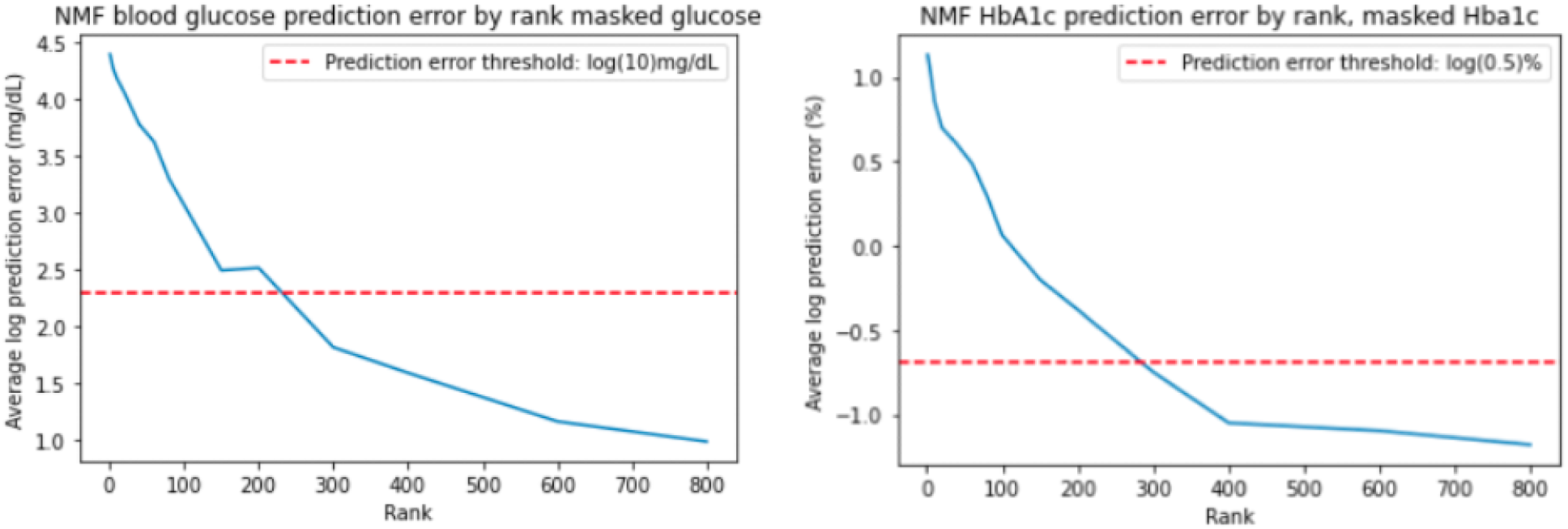
Prediction error vs. factorization rank for **blood glucose (left)** and **HbA1c (right)** using the masking approach. For each patient, the lab value of interest was masked during model training and then predicted from the patient’s latent representation derived from their remaining lab values.

#### Train Test Split Approach

In the second evaluation, we evaluated whether latent factors identified in one group of patients generalized to unseen patients. Figure 2 shows the prediction error across different latent dimensions for blood glucose and HbA1c, respectively. A minimum rank of ~250 was sufficient to predict blood glucose and minimum rank of ~300 was sufficient for HbA1c.

**Figure 2:**
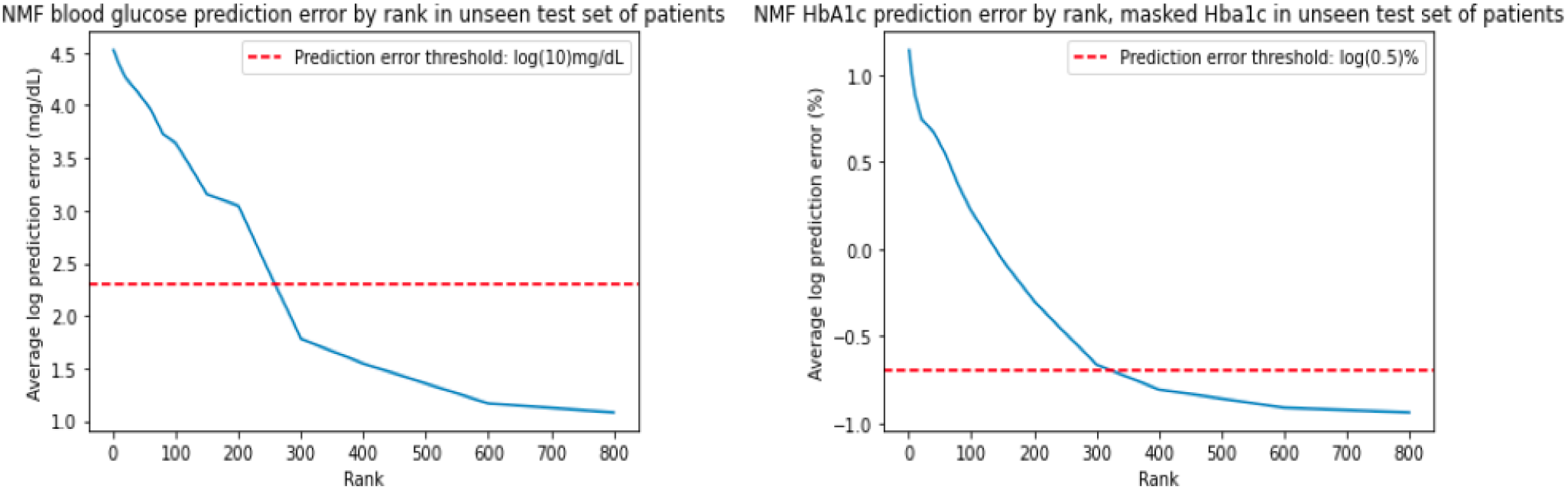
Prediction error vs. factorization rank for **blood glucose (left)** and **HbA1c (right)** using the train-test split approach. The model was trained on a subset of patients and then used to predict lab values for unseen patients, demonstrating cross-patient generalizability of the latent representations.

These results indicate that a relatively low-rank representation ~ 17 − 21% of the full lab dataset retains sufficient information to recover clinically meaningful outcomes, both within and across patients.

## 5 Discussion

Our analysis demonstrates that high-dimensional lab data from patients with type II diabetes can be effectively represented in a significantly lower-dimensional space while preserving clinically relevant information. Using NMF, we found that a rank of approximately 230-300 was sufficient to accurately reconstruct key diabetes management values—blood glucose and HbA1c—from a dataset with 1,418 laboratory features. This represents a dimensional reduction of approximately 80-85%, suggesting redundancy in clinical lab data.

The results from the masking approach approach indicates strong correlations between standard diabetes markers and other lab values, likely reflecting the systemic physiological changes associated with diabetes. Similarly, in the train-test split approach, which evaluated how well latent factors identified in one patient cohort generalized to unseen patients, accurate predictions were achieved at ranks of 250 for glucose and 300 for HbA1c. The modest increase in required dimensionality for the train-test approach (approximately 9% higher for glucose and 7% for HbA1c) suggests that the latent structure of laboratory data is relatively consistent across patients with type II diabetes.

### 5.1 Limitations

This study established the feasibility of identifying low-dimensional representations of lab data that preserve clinically relevant information in type II diabetes. However, several limitations exist. Our analysis was restricted to lab data from patients with type II diabetes at a single institution, potentially limiting generalizability to other clinical contexts or healthcare systems. Second, by using only the most recent laboratory values we discarded temporal data which likely captures valuable information. Third, we focused solely on lab data ignoring modalities such as diagnoses, medications, procedures, and clinical notes. Finally, we did not fully explore the clinical interpretation of identified latent dimensions.

### 5.2 Future directions

These limitations suggest promising avenues for future research. First, extending this framework to other diseases areas and prediction targets would validate the intrinsic dimension estimated here. Second, integrating multiple data modalities through methods like non-negative tensor factorization could provide more comprehensive patient representations. Third, developing methods to incorporate temporal patterns in laboratory measurements could capture disease progression dynamics. Finally, applying these latent representations to downstream clinical tasks such as risk prediction, patient stratification, or treatment response modeling would further demonstrate their utility in real-world clinical settings.

## Data Availability

All data produced in the present work are contained in the manuscript. Source medical record data are not available.

